# Identification of an ANCA-Associated Vasculitis Cohort Using Deep Learning and Electronic Health Records

**DOI:** 10.1101/2024.06.09.24308603

**Authors:** Liqin Wang, John Novoa-Laurentiev, Claire Cook, Shruthi Srivatsan, Yining Hua, Jie Yang, Eli Miloslavsky, Hyon K. Choi, Li Zhou, Zachary S. Wallace

## Abstract

**Background:** ANCA-associated vasculitis (AAV) is a rare but serious disease. Traditional case-identification methods using claims data can be time-intensive and may miss important subgroups. We hypothesized that a deep learning model analyzing electronic health records (EHR) can more accurately identify AAV cases.

**Methods:** We examined the Mass General Brigham (MGB) repository of clinical documentation from 12/1/1979 to 5/11/2021, using expert-curated keywords and ICD codes to identify a large cohort of potential AAV cases. Three labeled datasets (I, II, III) were created, each containing note sections. We trained and evaluated a range of machine learning and deep learning algorithms for note-level classification, using metrics like positive predictive value (PPV), sensitivity, F-score, area under the receiver operating characteristic curve (AUROC), and area under the precision and recall curve (AUPRC). The deep learning model was further evaluated for its ability to classify AAV cases at the patient-level, compared with rule-based algorithms in 2,000 randomly chosen samples.

**Results:** Datasets I, II, and III comprised 6,000, 3,008, and 7,500 note sections, respectively. Deep learning achieved the highest AUROC in all three datasets, with scores of 0.983, 0.991, and 0.991. The deep learning approach also had among the highest PPVs across the three datasets (0.941, 0.954, and 0.800, respectively). In a test cohort of 2,000 cases, the deep learning model achieved a PPV of 0.262 and an estimated sensitivity of 0.975. Compared to the best rule-based algorithm, the deep learning model identified six additional AAV cases, representing 13% of the total.

**Conclusion:** The deep learning model effectively classifies clinical note sections for AAV diagnosis. Its application to EHR notes can potentially uncover additional cases missed by traditional rule-based methods.

**SIGNIFICANCE AND INNOVATIONS:**

– Traditional approaches to identifying AAV cases for research have relied on registries assembled through clinical care and/or on billing codes which may miss important subgroups.
– Unstructured data entered as free text by clinicians document a patient’s diagnosis, symptoms, manifestations, and other features of their condition which may be useful for identifying AAV cases
– We found that a deep learning approach can classify notes as being indicative of AAV and, when applied at the case level, identifies more cases with AAV than rule-based algorithms.

## INTRODUCTION

Anti-neutrophil cytoplasmic autoantibody (ANCA)-associated vasculitis (AAV) is a rare immune-mediated inflammatory disease associated with substantial morbidity, mortality, and resource utilization.^1, 2^ The disease presents in patients with heterogeneous manifestations (e.g., glomerulonephritis, sinusitis, skin rash, pulmonary nodules) to clinicians from a variety of specialties (e.g., rheumatology, nephrology, otolaryngology, intensive care) across the care spectrum within healthcare systems (e.g., community hospital, tertiary care hospital, outpatient clinic, emergency departments). AAV case identification in electronic health records (EHR), an increasingly important source for epidemiologic research, is limited by a lack of well-performing methods to identify cases.

To enable outcomes and comparative effectiveness studies using large, phenotypically diverse cohorts from big data, a novel AAV case-finding algorithm is needed. Previous studies have demonstrated that rule-based algorithms relying on ICD-9 codes for AAV case identification have poor performance, partly because there is no specific ICD-9 code for microscopic polyangiitis (MPA), a subtype of AAV, and many MPA patients may be miscoded using less specific ICD-9 codes. Additionally, the previously developed algorithms, which require a positive ANCA test result or the use of multiple ICD codes, may exclude important and informative subgroups of patients, including ANCA-negative granulomatosis with polyangiitis (GPA) and those who die soon after diagnosis from severe disease or complications. The performance of algorithms that incorporate ICD-10 codes has not been previously assessed.

In addition to billing code data and test results, EHR data include unstructured data entered as free text by clinicians documenting a patient’s diagnosis, symptoms, manifestations, and other features of their condition. We have previously demonstrated that these notes can be leveraged to characterize the temporal course of AAV.^3^ Other studies have suggested that unstructured data can enhance the performance of case-finding algorithms for other conditions, but this remains underexplored for prototypic rare conditions like AAV.^4–7^ Here, we hypothesized that machine learning methods could be utilized to develop case-finding algorithms that accurately identify AAV patients and that these algorithms would outperform or produce more phenotypically diverse cohorts than rule-based algorithms.

## MATERIALS AND METHODS

### Overview

This study was conducted at Mass General Brigham (MGB), a large, integrated healthcare delivery system in the Greater Boston area, Massachusetts. We used data from MGB’s research patient data registry (RPDR). The study was approved by the MGB’s Institutional Review Board (IRB number: 2016P000633). **Figure 1** illustrates the overall process for the development of the AAV case-finding algorithms. We first created a screening cohort of potential AAV cases. We then created three labeled datasets from three cohorts for the development and evaluation of multiple machine learning models for AAV case identification from unstructured clinical notes. The deep learning model was further deployed to identify AAV patients from a random sub-cohort of patients. The performance of this model was compared against rule-based approaches.

**Figure 1.**
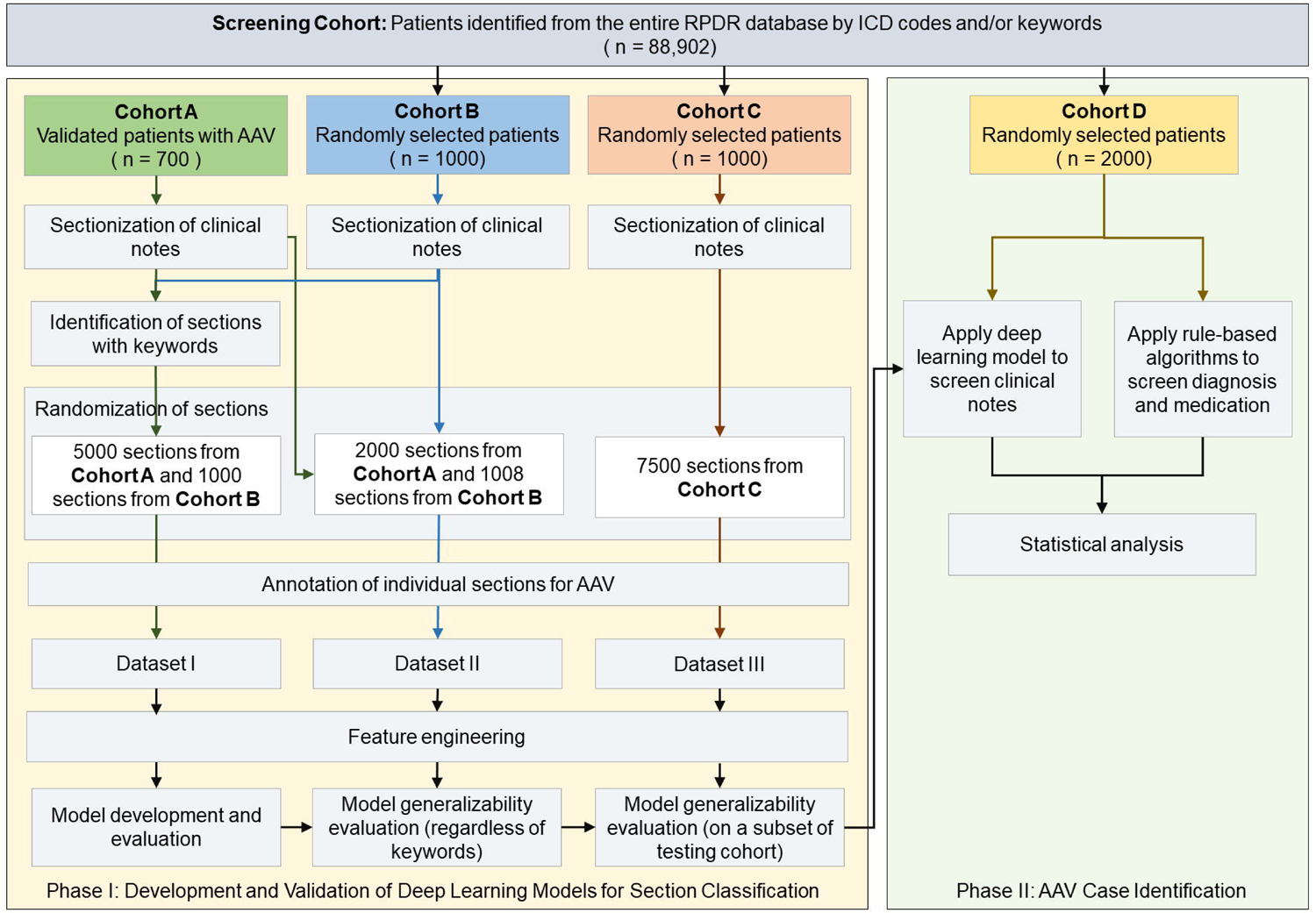
Two-Phase Process for Identifying ANCA-Associated Vasculitis Cases. Phase 1 entails dataset creation along with the training and evaluation of models. Phase 2 compared the performance of the deep learning model with two rule-based algorithms.

**Figure 2.**
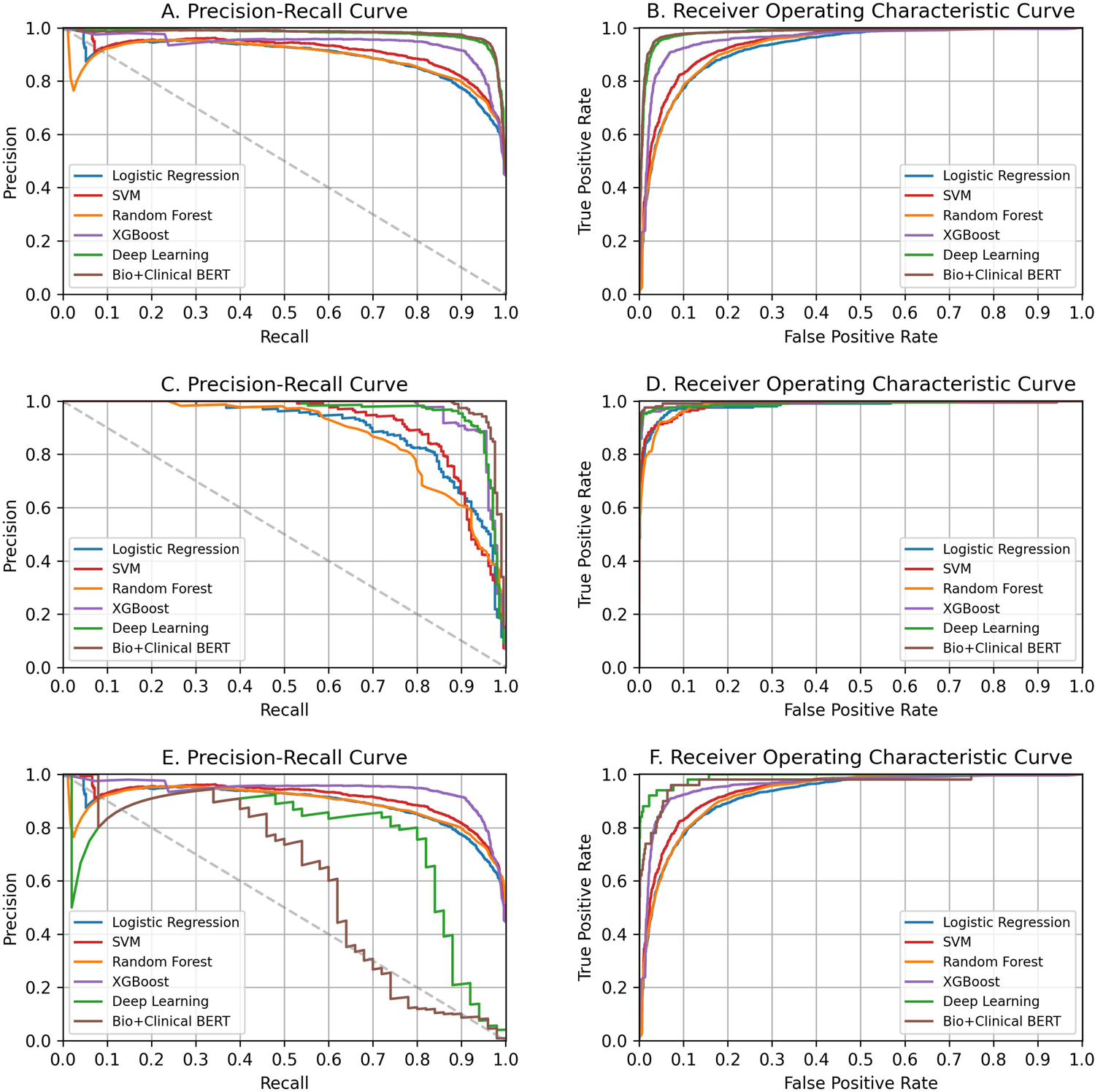
Performance of the Machine Learning and Deep Learning Algorithms on Datasets I, II, and III. A. Precision-recall curves for Dataset I. B. Receiver operating characteristic (ROC) curves for Dataset I. C. Precision-recall curves for Dataset II. D. ROC curves for Dataset II. E. Precision-recall curves for Dataset III. F. ROC curves for Dataset III.

### Study Cohorts

The screening cohort was constructed using all data from RPDR, spanning its inception on December 1, 1979, through May 11, 2021. We identified potential AAV patients based on the presence of at least one AAV-related ICD code in a diagnosis field or the use of a keyword in clinical notes. **Supplemental Table 1** lists keywords selected by the subject-matter experts (EM and ZSW). To develop and evaluate the performance of machine learning algorithms for AAV case identification, we used three distinct cohorts. Cohort A comprised 700 patients with confirmed AAV, as they were previously identified as part of the MGB AAV Cohort between January 01, 2002, and December 31, 2019.^8^ Cohorts B and C were random samples of 1000 patients each from the screening cohort.

### Processing of Clinical Notes

For each study cohort, we extracted all available clinical notes included in RPDR database any time before July 23, 2021. These notes encompassed visit notes, progress notes, ambulatory notes, history and physical exam notes, and discharge summaries. Clinical notes contain rich information, such as clinical manifestations, physical exams, and differential diagnoses, which are useful to determine whether patient does or does not have AAV. However, due to the voluminous number of notes that each patient accumulates over years of interaction with a healthcare system, the model could miss true signals when handling large datasets. Thus, we processed each note into smaller sections and then applied the models being tested to evaluate their ability to predict whether or not the text refers to a diagnosis of AAV. To split the notes into sections, we used Medical Text Extraction, Reasoning, and Mapping System (MTERMS), an in-house developed natural language processing (NLP) system.^9^

### Definition of AAV Classification Task

We approached the identification of AAV as a classification task, training models for binary classification of note sections as pertaining to AAV. Throughout model development, we assessed and compared the effectiveness of these models at the note section level. In practical applications for patient identification, a patient was classified as having AAV if any of their note sections were predicted positive, suggesting an AAV diagnosis. Conversely, a patient was deemed not to have AAV if all their note sections were predicted negative.

### Development of Labeled Datasets

We developed three labeled datasets to train, test, and compare multiple machine learning algorithms. Datasets I and II contained note sections that were derived from the same population, i.e., patients with validated AAV (Cohort A) as well as patients with possible AAV (Cohort B). To increase the positive case density in Dataset I, we applied a list of expert-curated keywords to filter for sections that likely contain references to a diagnosis of AAV or the presence of AAV manifestations (**Supplemental Table 2**). To assess the generalizability of the trained models to note sections, regardless of keyword presence, we established Dataset II by randomly selecting note sections from those not included in Dataset I. Specifically, Dataset I included 5,000 sections from cohort A and 1,000 sections from cohort B, all mentioning specific keywords. Dataset II contains 2,000 sections from cohort A and 1,000 sections from cohort B, selected randomly without considering keyword references. Dataset III consists of 7,500 note sections randomly selected from Cohort C, a subset of the screening cohort, to evaluate model performance in identifying potential AAV patients.

### Dataset Annotation

Two subject matter experts (ZSW and CC) labeled each selected section from clinical notes for whether it indicated that the diagnosis of AAV was present. Cases were classified as AAV based on a prior algorithm for identifying AAV in epidemiologic studies.^10^ In cases where there was limited data available to apply this algorithm, we classified a case as AAV if the treating provider and both chart reviewers agreed on the classification of the case as AAV. Patients with eosinophilic granulomatosis with polyangiitis (EGPA) were classified as negative. Although EGPA is a specific type of AAV, it presents a different etiology, pathology, and clinical features compared to other AAV types, such as GPA and MPA.^11^ The annotators first individually labeled 100 sections and any conflicts were resolved by consensus. Then in the second dataset of 100 sections, two annotators achieved near-perfect agreement with a Cohen’s kappa of 0.897. The remaining note sections were each annotated by one of the annotators. Any cases for which labeling was uncertain were resolved through consensus by ZSW and CC.

### Model Development

We first implemented four basic statistical machine learning algorithms, including logistic regression, random forest, support vector machine (SVM), and XGBoost.^12^ The note sections were processed into n-grams (where n=1). Each section was converted into term frequency-inverse document frequency vectors based on n-grams. The algorithms were trained and tested with 5-fold cross validation using the Dataset I.

We also implemented a hierarchical attention-based deep neural network, which includes a convolutional neural network for handling word variations (e.g., plural, misspelling), a recurrent neural network for handling context (e.g., negation), and attention layers for interpreting predictions. We chose this algorithm as it was previously proved to be effective in allergic reaction detection from hospital safety reports^5^ and cognitive decline detection from clinical notes.^4^ When implementing this deep learning algorithm, each note section was treated as a sequence of tokens, and individual words were represented by word embedding. We used pre-trained word embedding, named BioWordVec, which is an open set of biomedical word vectors that integrated biomedical text with Medical Subject Headings (MeSH) using the fastText model.^13^

Additionally, we implemented BioClinicalBERT, a domain-specific Bidirectional Encoder Representation from Transformers (BERT) model,^14^ on the labeled dataset. BERT is one of the most widely-used deep contextualized language models, achieving state-of-the-art performance on various NLP tasks, including named entity recognition, sentiment analysis, and question answering. We previously leveraged this algorithm to identify patient gender identity in the EHR.^7^

### Evaluation for Model Generalizability

To evaluate the generalizability of the models to the dataset regardless of keywords, we applied the models trained in Dataset I to Dataset II. To evaluate the generalizability of the models to the screening cohort, we applied the models trained in Datasets I and II to Dataset III and reported the models’ performance.

### Comparison of machine learning-based models with rule-based approaches

The assess the feasibility of applying the deep learning model to identify AAV cases, we compared its efficacy with that of two rule-based algorithms derived from administrative claims data. Specifically, Rule 1 identifies patients who have any ICD-9 or ICD-10 codes documented on at least two separated occasions. Rule 2 identifies patients with any ICD-9 or ICD-10 codes recorded on at least two separated dates, and who also received an AAV medication within a 6-month window (±6 months) of the first ICD code recorded. For the deep learning model, we used the optimal model to analyze the clinical notes of Cohort D. The highest section-level prediction probability was considered as the patient-level model prediction probability. Patients with a predicted probability of 1 were classified as positive for AAV.

After identifying potential AAV cases using either the rule-based or the deep learning model, we conducted a manual review of the EHR for these cases to pinpoint true positive cases. From the cases not deemed to be AAV (i.e., those not identified by the rules or the models), we randomly selected a subset (n=100) for manual chart review to assess the false negative rate.

### Statistical Analysis

We evaluated four statistical machine learning models, one deep learning model, and a large language model for AAV case detection from clinical notes. Performance was assessed based on the area under the receiver operating characteristic curve (AUROC), the area under the precision-recall curve (AUPRC), positive predictive value (PPV), sensitivity, and F-1 score which accounts for both precision and recall by taking the harmonic mean. Both the AUROC and AUPRC were computed using the scikit-learn Python library (scikit-learn Developers). We estimated the 95% confidence intervals (CI) using 2,000 bootstrap iterations (Python, version 3.7; Python Software Foundation). To compare the rule-based approaches with the top-performing AAV case identification model, we computed the PPVs and sensitivities of all methods in Cohort D. Here the total number of true positive patients was determined by adding those identified by both the rule-based approaches and the top-performing AAV case identification model.

## RESULTS

Cohort A, termed the 2002-2019 MGB AAV Cohort, included 700 PR3- or MPO-ANCA+ AAV patients. From these patients, 134,506 notes were extracted from the RPDR, which included progress notes, ambulatory notes, and discharge summaries. These notes were further split into 1,927,286 sections, of which 320,038 contained keywords. Dataset I comprised 6,000 note sections, representing 5,765 notes from 1,638 patients (968 [59.1%] female). Dataset II contained 3,008 sections from 2,970 notes, representing 1,501 patients (885 [60.0%] female). Dataset III included 7,500 sections, representing 5,429 notes from 1,000 patients (568 [56.8%] female) (**Table 1**). In Dataset I, evidence of AAV was present in 2,669 sections (44.5%). Dataset II had 206 (6.8%) sections positive for AAV. Out of the 3,008 sections in Dataset II, 457 contained at least one keyword, with 203 (44.4%) of these containing evidence of AAV. Dataset III had 50 (0.67%) AAV-positive sections, and, of the 7,500 sections, 219 (2.92%) had one or more keywords. Of those with keywords, 45 (20.5%) contained evidence of AAV.

**Table 1.** Characteristics of the datasets I, II and III for the development and validation of models for identifying evidence of ANCA vasculitis.

| Characteristic | Dataset I | Dataset II | Dataset III |
| --- | --- | --- | --- |
| Sections, n | 6,000 | 3,008 | 7,500 |
| Notes, n | 5,765 | 2,970 | 5,429 |
| Character length per section, mean (SD) | 839 (793) | 443 (566) | 410 (551) |
| Unique patients, n | 1,638 | 1,501 | 1,000 |
| Female, n (%) | 968 (59.1) | 885 (60.0) | 568 (56.8) |
| Keyword present, n (%) | 6,000 (100) | 457 (15.2) | 219 (2.9) |
| Sections consistent with a diagnosis of AAV, n (%) | 2,669 (44.5) | 206 (6.8) | 50 (0.67) |
| Sections consistent with AAV diagnosis and containing a keyword of interest, n (%) | 2,669 (44.5) | 203 (44.4) | 45 (20.5) |

Abbreviations: AAV, ANCA-associated vasculitis; SD, standard deviation

The performance of the five models in each dataset is outlined in **Table 2**. The hierarchical attention-based deep learning model demonstrated the best performance on Dataset I during cross-validation, significantly outperforming other models, with an AUROC of 0.983 (95% CI, 0.980-0.986) and an AUPRC of 0.977 (95% CI, 0.972-0.982). Compared to the deep learning model, Bio_ClinicalBERT had slightly worse performance in AUROC and AUPRC in Dataset I; however, it achieved better results in precision, recall, and F-1 score.

**Table 2.** Performance of four machine learning models for detecting AAV from clinical notes.

| Model | AUROC (95% CI) | AUPRC (95% CI) | PPV | Sensitivity | F-1 Score |
| --- | --- | --- | --- | --- | --- |
| **Dataset I (6,000 Note Sections)** |  |  |  |  |  |
| Logistic Regression | 0.923 (0.916-0.930) | 0.892 (0.879-0.904) | 0.857 | 0.777 | 0.815 |
| Random Forest | 0.929 (0.922-0.935) | 0.888 (0.874-0.902) | 0.808 | 0.886 | 0.845 |
| SVM | 0.938 (0.932-0.944) | 0.912 (0.901-0.923) | 0.848 | 0.862 | 0.855 |
| XGBoost | 0.957 (0.952-0.962) | 0.939 (0.929-0.948) | 0.916 | 0.896 | 0.906 |
| Bio_ClinicalBERT | 0.957 (0.945-0.968) | 0.962 (0.950- 0.972) | **0.947** | **0.956** | **0.952** |
| Deep Learning | **0.983 (0.980-0.986)** | **0.977 (0.972-0.982)** | 0.941 | 0.951 | 0.946 |
| **Dataset II (3,008 Note Sections)** |  |  |  |  |  |
| Logistic Regression | 0.981 (0.969-0.991) | 0.893 (0.858-0.925) | 0.709 | 0.874 | 0.783 |
| Random Forest | 0.983 (0.976-0.989) | 0.871 (0.832-0.904) | 0.528 | 0.922 | 0.671 |
| SVM | 0.983 (0.971-0.991) | 0.910 (0.880-0.939) | 0.645 | 0.908 | 0.754 |
| XGBoost | 0.990 (0.981-0.997) | **0.963 (0.941-0.981)** | 0.886 | 0.947 | 0.915 |
| Bio_ClinicalBERT | 0.981 (0.967-0.992) | 0.954 (0.933-0.973) | 0.939 | **0.966** | **0.952** |
| Deep Learning | **0.991 (0.981-0.997)** | 0.962 (0.941-0.980) | **0.954** | 0.898 | 0.925 |
| **Dataset III (7,500 Note Sections)** |  |  |  |  |  |
| Logistic Regression | 0.940 (0.906-0.967) | 0.390 (0.259-0.534) | 0.109 | 0.720 | 0.189 |
| Random Forest | 0.975 (0.962-0.986) | 0.392 (0.258-0.529) | 0.064 | 0.900 | 0.120 |
| SVM | 0.907 (0.861-0.948) | 0.402 (0.264-0.538) | 0.080 | 0.700 | 0.143 |
| XGBoost | 0.982 (0.955-0.996) | 0.490 (0.359-0.633) | 0.320 | 0.800 | 0.457 |
| Bio_ClinicalBERT | 0.857 (0.792-0.917) | 0.570 (0.473-0.659) | 0.419 | 0.720 | 0.529 |
| Deep Learning | **0.991 (0.982-0.998)** | **0.760 (0.620-0.885)** | **0.800** | **0.800** | **0.800** |

Abbreviations: SVM, support vector machine; AUROC, area under the receiver operating characteristic curve; AUPRC, the area under the precision-recall curve; PPV, positive predictive value. Bold highlights the best performing model according to each measure.

Overall, all the models generalized well to Dataset II. The deep learning model exhibited an AUROC of 0.991 (95% CI, 0.981-0.997) and an AUPRC of 0.962 (95% CI, 0.941-0.980), with a 0.015 drop in AUPRC compared to its performance in Dataset I. Notably, in Dataset II and among all the models, the XGBoost model achieved the best performance in AUPRC, though the difference was not statistically significant, and Bio_ClinicalBERT achieved the best sensitivity and F-1 score.

In Dataset III, the deep learning model outperformed other algorithms in all metrics. Compared to its performance in Datasets I and II, it maintained a high AUROC of 0.991 (95% CI, 0.982-0.998); however, the AUPRC decreased to 0.760 (95% CI, 0.620-0.885). Both Bio_ClinicalBERT and XGBoost saw greater decrease in performance from Datasets I and II to Dataset III.

Among 2,000 patients from the screening cohort, Rule 1 identified 218 with two or more AAV-related ICD codes (**Table 3**). After excluding 12 patients due to insufficient information to ascertain AAV status, 40 (19.4%) were confirmed to have AAV. Rule 2 identified 52 patients meeting Rule 1 criteria and receiving a medication prescription within 6 months of the first ICD code; 11 (21.2%) had confirmed AAV. Among the 2,000 patients, 1,977 had clinical notes reviewed using the deep learning model, which predicted AAV in 177 patients with a probability of 1. After excluding 5 patients due to insufficient information, 45 (26.2%) patients were confirmed to have AAV.

**Table 3.**
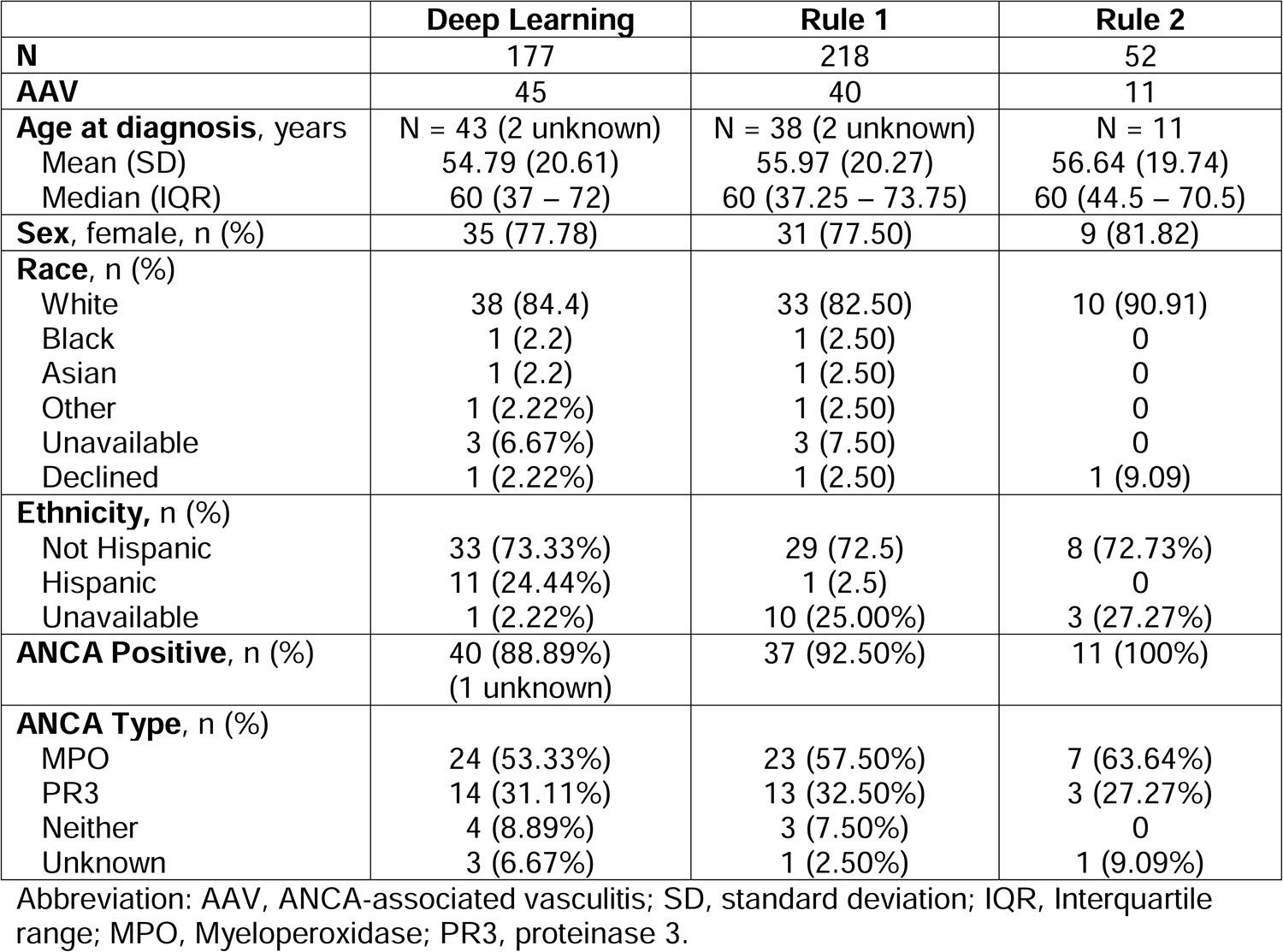
Characteristics of patients identified identified by the deep learning model and two rule-based algoirthms among 2,000 random sample of the screening cohort.

|  | **Deep Learning** | **Rule 1** | **Rule 2** |
| --- | --- | --- | --- |
| **N** | 177 | 218 | 52 |
| **AAV** | 45 | 40 | 11 |
| **Age at diagnosis, years** | N = 43 (2 unknown) | N = 38 (2 unknown) | N = 11 |
| Mean (SD) | 54.79 (20.61) | 55.97 (20.27) | 56.64 (19.74) |
| Median (IQR) | 60 (37 – 72) | 60 (37.25 – 73.75) | 60 (44.5 – 70.5) |
| **Sex, female, n (%)** | 35 (77.78) | 31 (77.50) | 9 (81.82) |
| **Race, n (%)** |  |  |  |
| White | 38 (84.4) | 33 (82.50) | 10 (90.91) |
| Black | 1 (2.2) | 1 (2.50) | 0 |
| Asian | 1 (2.2) | 1 (2.50) | 0 |
| Other | 1 (2.22%) | 1 (2.50) | 0 |
| Unavailable | 3 (6.67%) | 3 (7.50) | 0 |
| Declined | 1 (2.22%) | 1 (2.50) | 1 (9.09) |
| **Ethnicity, n (%)** |  |  |  |
| Not Hispanic | 33 (73.33%) | 29 (72.5) | 8 (72.73%) |
| Hispanic | 11 (24.44%) | 1 (2.5) | 0 |
| Unavailable | 1 (2.22%) | 10 (25.00%) | 3 (27.27%) |
| **ANCA Positive, n (%)** | 40 (88.89%)<br>(1 unknown) | 37 (92.50%) | 11 (100%) |
| **ANCA Type, n (%)** |  |  |  |
| MPO | 24 (53.33%) | 23 (57.50%) | 7 (63.64%) |
| PR3 | 14 (31.11%) | 13 (32.50%) | 3 (27.27%) |
| Neither | 4 (8.89%) | 3 (7.50%) | 0 |
| Unknown | 3 (6.67%) | 1 (2.50%) | 1 (9.09%) |

Abbreviation: AAV, ANCA-associated vasculitis; SD, standard deviation; IQR, Interquartile range; MPO, Myeloperoxidase; PR3, proteinase 3.

A review of 100 randomly selected cases, which were not predicted as AAV by either method, confirmed the absence of AAV cases. If both the rules and the deep learning algorithms identified all positive AAV cases among the 2000 cases reviewed, the total number of positive cases amounted to 46. The estimated sensitivities for the deep learning model, Rule 1, and Rule 2 were 97.5%, 87.0%, and 23.9%, respectively.

Significant differences were observed when comparing patients identified by the deep learning model versus rule-based algorithms (**Table 3**). The deep learning model identified a more ethnically diverse group (Hispanic: 24% vs 2.5% and 0%, respectively) and more ANCA-negative AAV cases. The deep learning model found six additional patients not identified by Rule 1, accounting for 13.0% of the positive cases, and Rule 1 found one not identified by the deep learning model.

### Error Analysis

We analyzed the sections of clinical notes which the deep learning model falsely predicted as consistent (false positive) with or not consistent (false negative) with AAV. Many of the false positive errors can be grouped into three categories.

1. Ambiguous terms related to AAV. Some terms that appear in clinical notes occasionally correspond to unrelated concepts with identical spelling. For example, the abbreviation “MPA” might denote microscopic polyangiitis, which is pertinent to AAV, or it could represent unrelated concepts like a multipurpose angiographic catheter or a Master of Public Administration degrees. Similarly, “GPA” can be used to abbreviate grade point average.

2, Hypothetical scenarios in notes. Prediction errors also arose in note sections described conjectural situations, such as guidelines of diagnosing AAV or potential medication side effects (e.g., risks of anti-thyroid medications).

3. Notes detailing family history. The model made false positive predictions on notes that mentioned a patient’s family member being diagnosed with AAV, even though the patient in question was not diagnosed.

False negative errors can be attributed to various representation of AAV-related keywords: 1. Dictation errors or misspellings. Some false negative cases contain typos of AAV-related terms or instances of terms transcribed incorrectly (e.g., “Wagner’s” instead of “Wegener’s”).

2. Combined terms. Certain terms related to AAV were mentioned as part of large tokens, which might not be recognized by the deep learning model. For instance, “ANCA+MPO+vasculitis” or “GPA/Wegener’s” were treated as distinct or unrelated compared to simpler terms like “ANCA+” or “GPA” or “Wegener’s”.

3. Rare variations of AAV-related terms, such as, “WEgeners”, “WEGENER’S” or “GRANULOMATOSIS”, which might not be well recognized by the deep learning model.

When assessing model performance in 2,000 random patients from our screening cohort, six patients were identified by the deep learning model but not by the rule-based algorithm. After reviewing their charts, there are two potential reasons for these patients were not captured by the rule-based algorithms. First, three patients were diagnosed with AAV at institutions external to MGB so ICD codes for AAV were not used in encounters in our healthcare system. Second, three patients had only one diagnosis code, which did not meet the criteria of our rule-based algorithm. One patient identified solely by the rule-based algorithm had positive ANCA pathology reports external to MGB, which weren’t included in the note screening. Available clinical notes lacked other specific features of AAV in this case.

## DISCUSSION

We found that a deep learning algorithm that integrated convolutional neural network, recurrent neural network, and an attention mechanism trained using a small set of keyword-identified, manually labeled note sections can be accurate and useful for identifying a rare disease like AAV in a large cohort. The model performance in dataset I showed its great capacity for detecting relevant signals from free-text narratives to make accurate predictions. The model was generalizable to notes, regardless of the presence of keywords. When applied to notes of patients from a large screening cohort for AAV case identification, the deep learning model out-performed the traditional rule-based algorithms which rely on ICD codes with or without medication prescriptions.

In addition to assessing the performance of the deep learning model, we also evaluated the performance of rule-based algorithms using ICD codes and medication prescriptions in our healthcare system. This is the first study that incorporates ICD-10 codes into an assessment of performance of this rule-based algorithm. We found that these rule-based algorithms had a PPV worse than that of the deep-learning model and that incorporating medication prescriptions into the rule only slightly improved the PPV by 1.8%. In contrast, requiring a medication prescription significantly reduced sensitivity. These observations speak to the need for innovative approaches, such as deep learning, for developing new approaches for AAV case identification.

In comparing the deep learning approach with the ICD/medication-based rules, the former demonstrated higher sensitivity and PPV. Examining clinical notes proved beneficial in identifying additional cases, particularly those with a more remote history of AAV or those diagnosed with AAV outside the MGB system. It also addressed cases missed by the rule-based algorithm due to a limited number of ICD codes in the EHR. This will be helpful for identifying patients, for instance, who have severe disease and die during their initial admission for AAV. This is particularly crucial in rare diseases, where even a small increase in sample size and including patients with the most severe spectrum of disease can significantly impact studies. While clinical notes revealed only 6 additional cases in a sample of 2,000, after extrapolating these observations to the entire screening cohort (n=88,902) we suspect that the deep learning model could identify approximately 7,868 patients, with an estimated 2,000 of them having AAV. Compared with a rule-based algorithm approach, the deep learning algorithms could identify an additional 267 patients while reducing the need for extensive chart reviews by more than 1,823 patients.

Compared with rule-based algorithms, we found that the deep learning model more often identified patients of Hispanic background and those with ANCA-negative disease. Why the deep learning model may have yielded a cohort with greater ethnic diversity is unclear. One possibility has to do with differences in the way that ICD codes are used for billing between people of different racial or ethnic backgrounds or because of the way patients of different racial or ethnic backgrounds interact with the healthcare system. The ability of the deep learning model to identify a greater proportion of cases with ANCA-negative granulomatosis with polyangiitis is another strength of this approach. This population is often excluded from observational studies of AAV as well as clinical trials and the ability to identify them easily will facilitate research of this subgroup.

Our findings suggest that applying a deep learning model may have benefit regarding the efficiency of AAV cases identification. Rule-based approaches to AAV case finding which identify potential AAV cases through billing codes with or without medications typically necessitates a full chart review. In contrast, the deep learning model approach presents a significant advantage because once the model flags sections that are potentially related to AAV, only these specific sections typically require review, potentially reducing the need for comprehensive chart evaluation.

Our study has several strengths. First, it was conducted in a large healthcare system that includes both quaternary academic medical centers in addition to community hospitals, primary care and specialty clinics, as well as specialty hospitals (e.g., ear, nose throat hospital). Second, we applied four statistical machine learning models and assessed their performance in comparison to commonly used rules-based algorithms. Third, we assessed model performance using training and multiple validation datasets.

Despite these strengths, this study has certain limitations. First, we used data from a single healthcare system so the model was not evaluated for its generalizability using data from other institutions. This is an important next step in the development of deep learning models to identify AAV cases. Second, the deep learning model was learned from a small dataset, of which the vocabulary size might be relatively small. This might affect the performance of the model when applied to a dataset with a larger vocabulary size. Third, the current approach leveraged only clinical notes to identify potential AAV cases. It is possible that including other data sources for model learning, such as lab results, may improve the performance of algorithms for identifying AAV cases. Fourth, we noted a large decline in the PPV when we applied the deep learning model, which was trained and assessed at the level of sections from notes, to classify at the patient level. This decrease can be attributed to the aggregation of errors from multiple note sections per patient, which, when accumulated at the patient level, magnify the error rate.

Our findings highlight the potential role of deep learning models for identifying positive AAV cases from large screening cohorts. Moving forward, we intend to leverage our deep learning model to screen the entire cohort, anticipating the identification of over 2,000 AAV cases. A cohort of this size would be substantially larger than the current cohort assembled during a similar timeframe which includes fewer than 1,000 cases. Thus, this represents a significant opportunity to expand the current MGB AAV cohort. Furthermore, in the wake of the rise of sophisticated large language models, such as GPT-4, an intriguing avenue of research would be to compare the performance of our deep learning model with these state-of-the-art language models. The evolution of natural language processing tools offers promising opportunities to further enhance the accuracy and efficiency of clinical data mining and disease identification.

## CONCLUSION

This study is the first to show that a deep learning algorithm can efficiently and accurately identify cases of AAV, a prototypic rare condition, in part by only using unstructured EHR data. This approach has the potential to identify cases that may be overlooked if only using structured EHR data. This approach to case identification may improve the spectrum of disease captured for observational studies and reduce the time and resources often needed to review electronic health records. Future work will involve deploying the model to screen a broader cohort for potential AAV patients and assessing performance in other healthcare systems.

## Data Availability

All data produced in the present study are available upon reasonable request to the authors

## References

1. Kitching AR, Anders H-J, Basu N, Brouwer E, Gordon J, Jayne DR, et al. ANCA-associated vasculitis. Nature reviews Disease primers. 2020;6(1):71.

2. Tan JA, Dehghan N, Chen W, Xie H, Esdaile JM, Avina-Zubieta JA. Mortality in ANCA-associated vasculitis: ameta-analysis of observational studies. Annals of the rheumatic diseases. 2017;76(9):1566–74.

3. Wang L, Miloslavsky E, Stone JH, Choi HK, Zhou L, Wallace ZS. Topic modeling to characterize the natural history of ANCA-Associated vasculitis from clinical notes: A proof of concept study. Semin Arthritis Rheum. 2021;51(1):150–7. PMID: 33383291.

4. Wang L, Laurentiev J, Yang J, Lo YC, Amariglio RE, Blacker D, et al. Development and Validation of a Deep Learning Model for Earlier Detection of Cognitive Decline From Clinical Notes in Electronic Health Records. JAMA Netw Open. 2021;4(11):e2135174. PMID: 34792589.

5. Yang J, Wang L, Phadke NA, Wickner PG, Mancini CM, Blumenthal KG, et al. Development and Validation of a Deep Learning Model for Detection of Allergic Reactions Using Safety Event Reports Across Hospitals. JAMA Netw Open. 2020;3(11):e2022836. PMID: 33196805.

6. Shao Y, Zeng QT, Chen KK, Shutes-David A, Thielke SM, Tsuang DW. Detection of probable dementia cases in undiagnosed patients using structured and unstructured electronic health records. BMC Med Inform Decis Mak. 2019;19(1):128. PMID: 31288818.

7. Hua Y, Wang L, Nguyen V, Rieu-Werden M, McDowell A, Bates DW, et al. A deep learning approach for transgender and gender diverse patient identification in electronic health records. J Biomed Inform. 2023;147:104507. PMID: 37778672.

8. Wallace ZS, Fu X, Cook C, Ahola C, Williams Z, Doliner B, et al. Comparative Effectiveness of Rituximab-Versus Cyclophosphamide-Based Remission Induction Strategies in Antineutrophil Cytoplasmic Antibody-Associated Vasculitis for the Risk of Kidney Failure and Mortality. Arthritis Rheumatol. 2023;75(9):1599–607. PMID: 37011036.

9. Zhou L, Plasek JM, Mahoney LM, Karipineni N, Chang F, Yan X, et al. Using Medical Text Extraction, Reasoning and Mapping System (MTERMS) to process medication information in outpatient clinical notes. AMIA Annu Symp Proc. 2011;2011:1639–48. PMID: 22195230.

10. Watts R, Lane S, Hanslik T, Hauser T, Hellmich B, Koldingsnes W, et al. Development and validation of a consensus methodology for the classification of the ANCA-associated vasculitides and polyarteritis nodosa for epidemiological studies. Ann Rheum Dis. 2007;66(2):222–7. PMID: 16901958.

11. Kitching AR, Anders HJ, Basu N, Brouwer E, Gordon J, Jayne DR, et al. ANCA-associated vasculitis. Nat Rev Dis Primers. 2020;6(1):71. PMID: 32855422.

12. Chen T, Guestrin C, editors. Xgboost: A scalable tree boosting system. Proceedings of the 22nd acm sigkdd international conference on knowledge discovery and data mining; 2016.

13. Zhang Y, Chen Q, Yang Z, Lin H, Lu Z. BioWordVec, improving biomedical word embeddings with subword information and MeSH. Scientific data. 2019;6(1):52.

14. Devlin J, Chang M-W, Lee K, Toutanova K. Bert: Pre-training of deep bidirectional transformers for language understanding. arXiv preprint arXiv:181004805. 2018.

