## Supplemental Table 1 for "Identification of an ANCA-Associated Vasculitis Cohort Using Deep Learning and Electronic Health Records"

**Online Supplement**

**Supplemental Table 1.** ICD codes, medications, and keywords used to search for potential AAV cases.

| **ICD codes** | |
| --- | --- |
| **ICD-9 Codes** | 446.4 Granulomatosis with polyangiitis  446.0 Polyarteritis nodosa  447.6 Arteritis, unspecified  447.5 Necrosis of artery |
| **ICD-10 Codes** | I77.6 Arteritis, unspecified  M31.7 Microscopic polyangiitis  M31.3X Granulomatosis with polyangiitis |
| **Medications** | Rituximab, Rituximab-abbs, Truxima, Rituxan, Cytoxan, Cellcept, Methotrexate, Cyclophosphamide, Mycophenolate mofetil, Myfortic, Imuran, Azathioprine, Prednisone, Solumedrol, Methylprednisolone, Dexamethasone, Decadron, Prednisolone, and Mycophenolic acid |
| **Keywords** | ANCA  AAV  Wegener  Granulomatosis with polyangiitis  Anti-neutrophil cytoplasmic antibody  Myeloperoxidase  Proteinase  Microscopic polyangiitis  ANCA associated vasculitis  Granulomatosis |

**Supplemental Table 2**. Expert-curated keywords related to AAV or related manifestations.

**Manually Curated Keywords**

Wegener granulomatosis GPA MPA Microscopic polyangiitis ANCA-associated AAV Vasculitis ANCA Anti-neutrophil cytoplasmic antibody Proteinase Myeloperoxidase Glomerulonephritis RPGN Tracheal stenosis Subglottic Nasal crusting PR3-ANCA MPO-ANCA Diffuse alveolar hemorrhage RBC purpura Leukocytoclastic LCV Granulomatous Saddle nose deformity C-ANCA P-ANCA Vasculitic neuropathy MPO orbital pseudotumor Mononeuritis multiplex Destructive sinusitis Pachymeningitis Nephritic

Note: the keywords were case insensitive during the keyword-based search.
